# Protocol for the HIT-Stroke Trial 2 randomized controlled trial: Moderate-intensity exercise versus high-intensity interval training to determine the optimal training intensity for walking rehabilitation in chronic stroke

**DOI:** 10.1101/2025.07.30.25332470

**Authors:** Christina Garrity, Darcy S. Reisman, Sandra A. Billinger, Daria Pressler, Erin Wagner, Oluwole Awosika, Bria L. Bartsch, Amanda Briton-Carpenter, Daniel Carl, Amanda Engler, Jolene Foster, Jemma Kim, Kiersten McCartney, Alexandra Moores, Heidi Sucharew, Elizabeth Thompson, Katherine Walters, Emily Wasik, Henry Wright, Madison Yeazell, Pierce Boyne

**Affiliations:** Department of Rehabilitation, Exercise and Nutrition Sciences, College of Allied Health Sciences, University of Cincinnati, Cincinnati, OH, United States; Department of Physical Therapy, University of Delaware, 540 S, College Ave, Newark, DE, United States; Biomechanics and Movement Science Program, University of Delaware, 540 S. College Ave, Newark, DE, United States; Department of Neurology, School of Medicine, University of Kansas Medical Center, Kansas City, KS, United States; Department of Cell Biology and Integrative Physiology, School of Medicine, University of Kansas Medical Center, Kansas City, KS, United States; University of Kansas Alzheimer’s Research Disease Center, Fairway, KS, United States; Department of Physical Medicine and Rehabilitation, School of Medicine, University of Kansas Medical Center, Kansas City, KS, United States; Department of Emergency Medicine, College of Medicine, University of Cincinnati, Cincinnati, OH, United States

**Keywords:** Gait, Dose, Rehabilitation, Locomotion, Treadmill, Overground, Clinical Trial

## Abstract

**Background:** Current practice guidelines recommend moderate to vigorous intensity locomotor training to improve walking outcomes in chronic stroke. However, these intensities span a wide range, and the lack of specificity may lead to under-dosing or over-dosing of training intensity. Recent evidence indicates that vigorous intensity locomotor training improves walking outcomes significantly more than moderate intensity. Although, previous studies have not been powered to rule out the possibility of meaningful risk increases or negligible benefit with vigorous versus moderate intensity, nor have they been designed to compare sustained effects after training ends. In addition, small subgroup analyses have suggested that individuals with severe walking limitations (speed <0.4 m/s) may require vigorous training intensity to have meaningful benefit, but this has not been prospectively tested with a sufficient sample. The results of this study are expected to provide more specific guidance for optimizing locomotor training intensity and walking outcomes in chronic stroke.

**Methods:** In this single-blind, 3-site, randomized trial, 156 chronic (>6 months) stroke survivors will be allocated to 36 sessions (3 times a week for 12 weeks) of either high intensity interval or moderate intensity continuous locomotor training. Eligible participants have residual walking limitations from stroke and can walk without continuous physical assistance from another person. At least 52 participants will have severe baseline walking speed limitations. Outcomes are assessed at baseline, after 4 weeks, 8 weeks, 12 weeks (POST), and 3 months after completing training. The primary outcome is walking capacity (6-minute walk distance). Secondary outcomes include comfortable and fast gait speed, aerobic capacity, fatigue, balance confidence, quality of life, and motivation for exercise. Statistical analyses will compare outcome changes and adverse events between treatment groups, and will include subgrouping by walking limitation severity.

**Discussion:** This study will provide important new information to guide greater specificity and individualization of locomotor training intensity in chronic stroke.

**Trial Registration:** ClinicalTrials.gov NCT06268041; Registration Date: 2024-02-12

## BACKGROUND

Among the 9 million stroke survivors living in the United States^1^, 73% do not regain adequate walking speed and endurance (i.e. walking capacity) for normal daily activity.^2–4^ This limitation in walking capacity can be caused by both neurologic gait impairments from the stroke and aerobic deconditioning due to inactivity.^5,6^ Current rehabilitation practice guidelines recommend moderate to high intensity locomotor training to improve walking outcomes in chronic stroke.^6–8^ However, these intensity recommendations span a very wide range, with moderate intensity typically defined as 40-60% heart rate reserve (HRR) and high/vigorous intensity typically defined as 60-90% HRR, or nearing maximal work rate (e.g. training speed).^9,10^ This lack of specificity could lead to under- or over-dosing of locomotor training intensity.

Compared with moderate locomotor training intensity, vigorous intensity appears to elicit greater gains in walking capacity and/or aerobic fitness.^11–13^ For example, HIT-Stroke Trial 1 compared high-intensity interval training (HIIT) and moderate-intensity aerobic training (MAT); each involving up to 36 sessions of overground and treadmill walking over 12 weeks.^14^ HIIT improved 6-minute walk distance (6MWD) significantly more than MAT after 8 and 12 weeks of training, with increasing gains and between-group differences over time. Compared with MAT, HIIT also elicited significantly greater improvements in comfortable gait speed (CGS) and fastest gait speed (FGS) after 4, 8, and 12 weeks of training with greater improvement in fatigue at the 8-week time point. There were no serious adverse events related to study procedures and no significant between-group difference in adverse events.

Despite the promising results from this preliminary trial with 55 participants, more data is needed to clarify the relative benefits and risks of vigorous versus moderate intensity locomotor training. For example, the modest sample size of HIT-Stroke Trial 1 made it underpowered to detect all but large differences in adverse event risk between HIIT and MAT. The between group estimates of treatment effect were also relatively imprecise, with wide 95% confidence intervals (CI) that spanned both meaningful and trivial differences in walking capacity gains. Consequently, the results cannot rule out the possibility of meaningful increases in risk or negligible increases in benefit with vigorous versus moderate intensity locomotor training. Thus, a larger trial is needed to provide more conclusive evidence to guide intensity dosing for walking rehabilitation.

Additional evidence is also needed to clarify whether vigorous training intensity may be particularly important for individuals with severe baseline walking limitations (CGS < 0.4 m/s). In this lower prognosis subgroup, moderate intensity locomotor training has not shown meaningful changes in walking capacity.^14,15^ For example, Dean et al^15^ found non-significant walking capacity gains of only 17 meters (95% CI: −25, 59) with MAT in those with baseline CGS < 0.4 m/s (N=11). In the HIT-Stroke Trial 1,^14^ this subgroup had a non-significant walking capacity gain of only 10 meters with MAT (95% CI: −40, 60; N=7), but had a significant and meaningful gain of 55 meters with HIIT (95% CI: 15, 95; N=7). This suggests vigorous locomotor training may be required to generate meaningful walking capacity gains in those with severe baseline walking impairment. However, the small subgroup samples in these previous studies resulted in the imprecise confidence intervals, which cannot rule out meaningful gains with MAT or trivial gains with HIIT. Further, prior studies were not designed to test vigorous vs. moderate intensity comparisons within this subgroup. This warrants a trial with more participants who have severe baseline walking speed limitations.

Lastly, it is still unknown if the net gains in walking capacity from vigorous vs. moderate intensity locomotor training are robust enough to be sustained after training ends, since previous studies comparing these intensities have not included follow-up assessment. This implies that key performance measures (e.g. fatigue, quality of life, daily walking activity) tested in the post-intervention assessment were likely confounded by acute training effects (e.g. fatigue). Thus, a trial with a longer term follow-up assessment is needed to evaluate retention effects and provide more valid assessment of some relevant measures.

Given the gaps identified above, the present study aims to: (1) determine the optimal training intensity to safely elicit meaningful and sustained improvements in walking capacity in chronic stroke; and (2) assess whether vigorous training intensity is particularly critical for patients with severe walking limitations (CGS <0.4 m/s).

## METHODS

This HIT-Stroke Trial 2 protocol was adapted from HIT-Stroke Trial 1^14^ as detailed in Table 1.

**Table 1.**
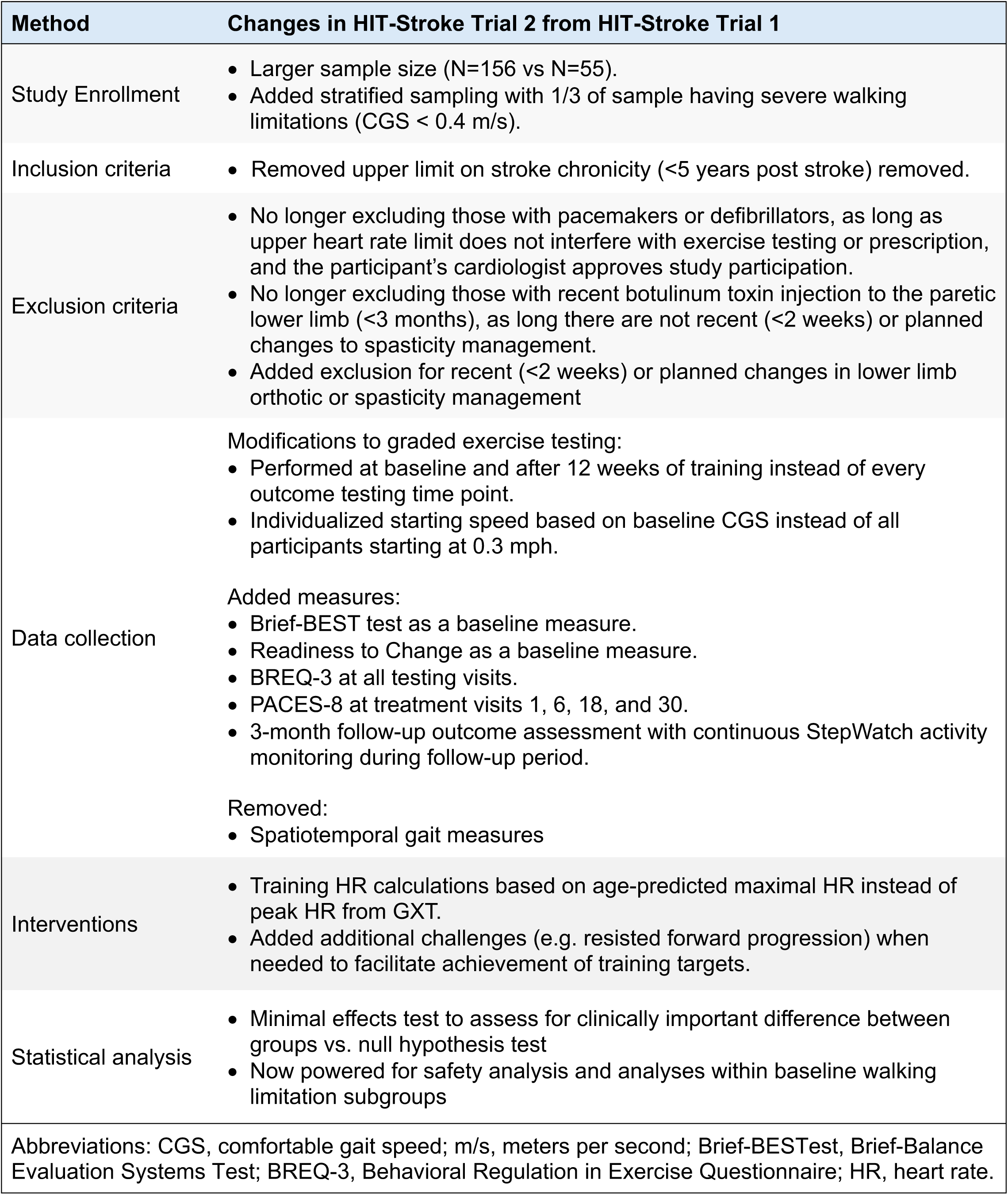
Present study methods that differ from HIT-Stroke Trial 1.

### Trial design

This is a single-blind, 3-site, randomized, active-control trial comparing HIIT and MAT. Participants undergo screening and pre-testing (PRE) to determine eligibility and obtain baseline data. Eligible participants are then randomized into either the MAT or HIIT protocol. Both protocols include up to 36 sessions with a goal of 3x/week for 12 weeks. Participants undergo outcome testing by a blinded physical therapist (PT) at baseline (PRE), after 4 weeks of training (4-WK), after 8 weeks of training (8-WK), at the end of the 12-week training protocol (POST), and 3 months after the completion of the last training session (3moPOST). Figure 1 shows the study flow diagram and Table 2 shows the outcomes of interest with prespecified time points.

**Figure 1.**
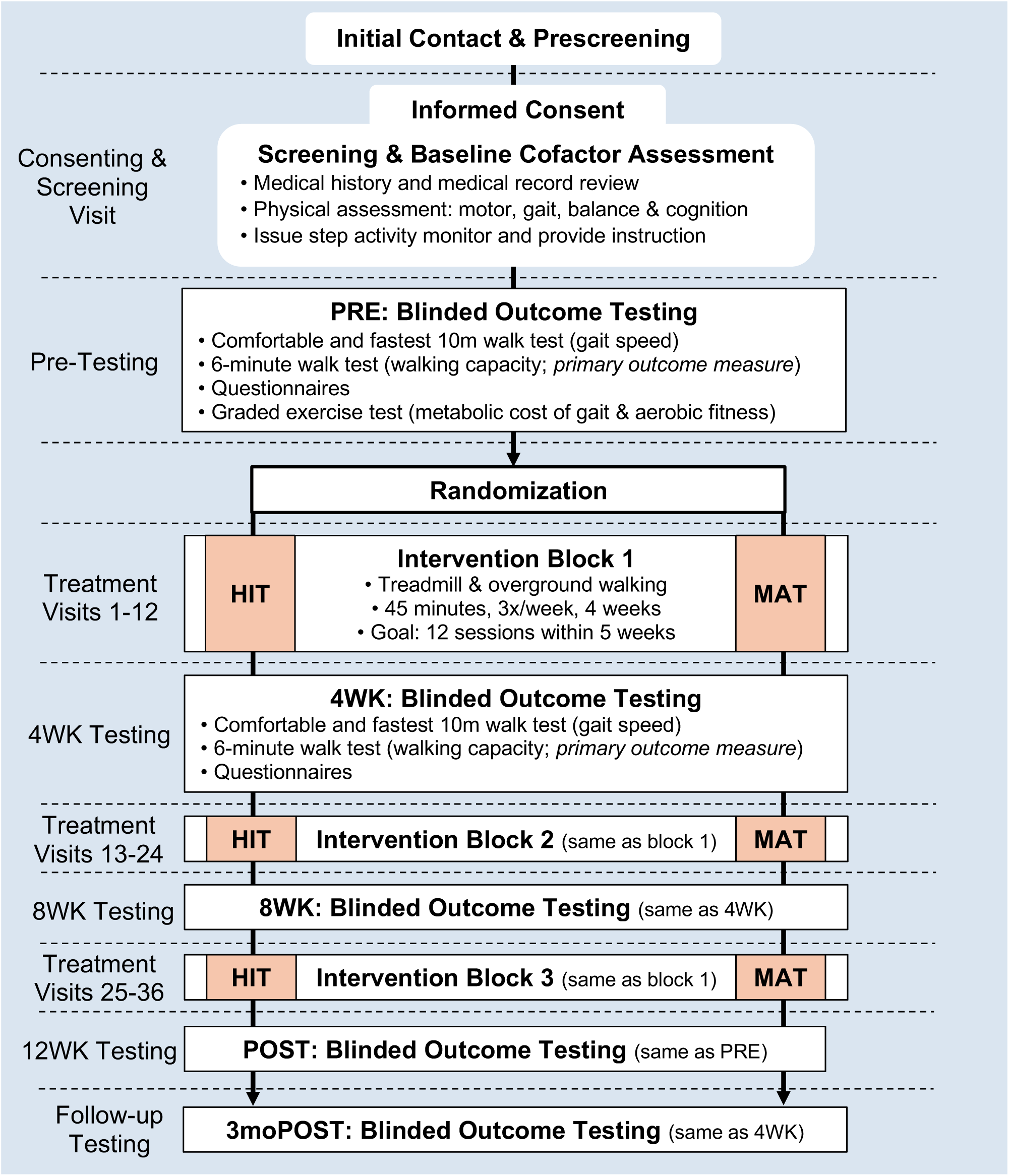
Participant flow diagram from initial contact through study completion.

**Table 2.**
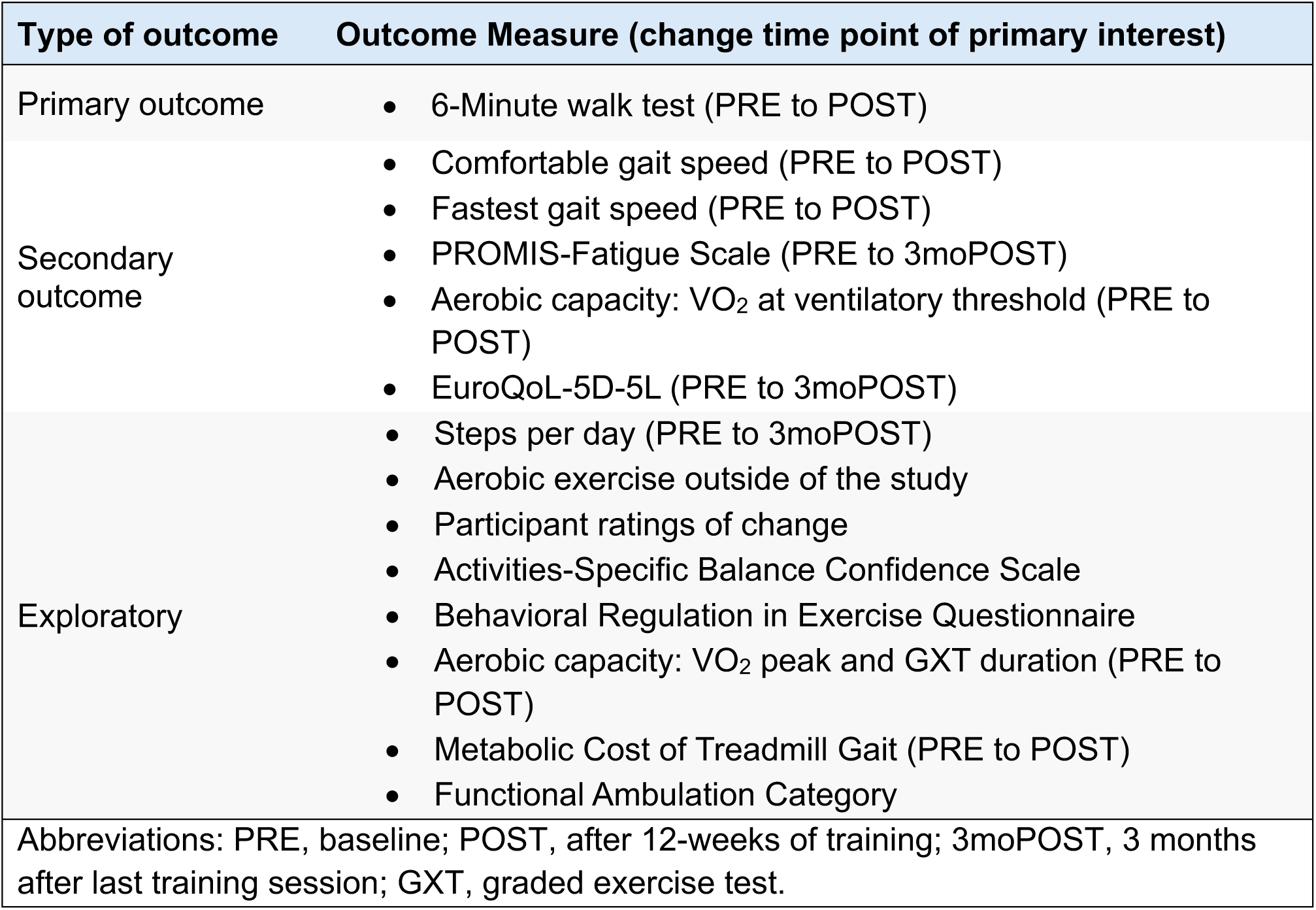
Outcomes of interest with prespecified time points for change measurement.

| Type of outcome | Outcome Measure (change time point of primary interest) |
| --- | --- |
| Primary outcome | • 6-Minute walk test (PRE to POST) |
| Secondary outcome | • Comfortable gait speed (PRE to POST)<br>• Fastest gait speed (PRE to POST)<br>• PROMIS-Fatigue Scale (PRE to 3moPOST)<br>• Aerobic capacity: VO<sub>2</sub> at ventilatory threshold (PRE to POST)<br>• EuroQoL-5D-5L (PRE to 3moPOST) |
| Exploratory | • Steps per day (PRE to 3moPOST)<br>• Aerobic exercise outside of the study<br>• Participant ratings of change<br>• Activities-Specific Balance Confidence Scale<br>• Behavioral Regulation in Exercise Questionnaire<br>• Aerobic capacity: VO<sub>2</sub> peak and GXT duration (PRE to POST)<br>• Metabolic Cost of Treadmill Gait (PRE to POST)<br>• Functional Ambulation Category |
| Abbreviations: PRE, baseline; POST, after 12-weeks of training; 3moPOST, 3 months after last training session; GXT, graded exercise test. |  |

### Study enrollment

We plan to enroll 156 individuals >6 months post stroke, including at least 52 with severe baseline walking limitations, defined by CGS < 0.4 m/s. Participants are recruited across 3 sites: University of Cincinnati (UC), University of Delaware (UD), and University of Kansas Medical Center (KUMC). A variety of recruitment sources are used including existing clinical databases, local physical therapists and physicians, local stroke support groups, and advertisements.

### Screening process

A member of the research team provides a brief overview of the study and describes potential risks and benefits of participation to determine initial interest via a phone call or email. Individuals who express interest answer pre-screening questions to determine initial eligibility. Potential participants who meet initial eligibility criteria are then scheduled for an in-person screening visit. Participants complete the informed consent process prior to in-person screening.

#### Inclusion and exclusion criteria

Inclusion criteria are: (1) Age 30-85 years at time of consent; (2) Hemiparesis from ischemic and/or hemorrhagic stroke(s); (3) Most recent stroke for which the participant sought treatment at least 6 months prior to consent; (4) CGS <1.0 m /s on the 10-meter walk test; (5) Able to walk 10m over ground with assistive devices as needed and no continuous physical assistance from another person (guarding and intermittent assistance for loss of balance allowed); (6) Able to walk at least 3 minutes on the treadmill at ≥0.13 m/s (0.3 miles per hour [mph]); (7) Stable cardiovascular condition (American Heart Association class B, allowing for aerobic capacity <6 METs); (8) Able to communicate with investigators, follow a 2-step command and correctly answer consent comprehension questions.

Exclusion criteria are: (1) Exercise testing uninterpretable for ischemia or arrhythmia (e.g. resting electrocardiogram [ECG] abnormality that makes exercise ECG uninterpretable for ischemia and no other clinical testing from the past year available to rule out); (2) Evidence of significant arrhythmia or myocardial ischemia on treadmill ECG graded exercise test in the absence of recent (past year) more definitive clinical testing (e.g. stress nuclear imaging) with negative result; (3) Hospitalization for cardiac or pulmonary disease within past 3 months; (4) Implanted pacemaker or defibrillator with an upper heart rate limit that would interfere with exercise testing or prescription, or with unknown limit; (5) Significant ataxia or neglect (score of 2 on NIH stroke scale item 7 or 11); (6) Severe lower limb spasticity (Ashworth >2); (7) Known recent history (<3 months) of unstable substance abuse or unstable mental illness; (8) Major post-stroke depression (Patient Health Questionnaire ≥ 10) in the absence of depression management by a health care provider; (9) Currently participating in physical therapy or another interventional study targeting walking function; (10) Recent (<2 weeks) or planned changes in lower limb orthotic or spasticity management; (11) Foot drop or lower limb joint instability without adequate stabilizing device, as assessed by a PT; (12) Clinically significant neurologic disorder other than stroke; (13) Unable to walk outside the home prior to stroke; (14) Other significant medical condition likely to limit improvement or jeopardize safety as assessed by a PT (e.g. joint contracture, gait limited by pain); (15) Pregnancy; (16) Previous exposure to fast treadmill walking (>3 cumulative hours) in the past year.

### Testing visits

Eligible participants have 5 total testing visits performed by a therapist who is blinded to the intervention group. The PRE and POST outcome testing visits follow the same procedures and include gait testing, questionnaires, and a treadmill graded exercise testing (GXT). The 4-WK, 8-WK, and 3moPOST outcome testing visits only include gait testing and questionnaires.

#### Pre-testing visit

During the screening visit, height and weight are recorded, and medical history is taken to calculate the Charlson Comorbidity Index^16^ and the modified Functional Comorbidity Index.^17^ The Patient Health Questionnaire-9 is used to screen for depressive symptoms.^18^ Impairment and mobility testing includes ability to follow 2-step commands, Ashworth Scale^19^ for lower limb spasticity, ataxia and neglect items from NIH Stroke Scale^20^, walking-related pain, and comfortable and fast gait speed^21^ to determine study eligibility. Treadmill acclimation is performed to familiarize participants to walking on a treadmill. To determine eligibility, participants must walk at least 0.3 mph for 3 consecutive minutes with the goal of reaching or exceeding their fastest overground speed if able. A physical therapist observes the participant’s gait to screen for patterns that would place the participant at high risk for orthopedic injury during fast treadmill walking (e.g. severe ankle inversion, severe knee hyperextension). During acclimation, participants wear an overhead safety harness and heart rate (HR) is monitored using a Polar H10 heart rate monitor and maintained below 85% of age-predicted HRR.

Additional baseline tests are performed during the initial screening to characterize the study sample. These include:

- *Brief-Balance Evaluation Systems Test* – an 8-item balance measure assessing 6 balance domains.^22^
- *Lower extremity Fugl-Meyer Motor Assessment* – a stroke-specific measure of lower extremity motor control of fractionated movements and reflexes.^23^
- *Readiness to Change* – a 1-item questionnaire where participants select a statement that best describes their current and intended activity level.^24^

#### Assessments

##### Gait testing

All gait testing is performed using any orthotic and assistive device the participant most often uses during normal daily walking. A blinded tester administers each of the following measures during the PRE, 4WK, 8WK, POST, and 3moPOST testing visit:

- *Comfortable and fast gait speed* – measured using the 10-meter walk test, allowing an untimed 2-meter acceleration distance, followed by 10-meter of timed walking, and an untimed 2-meter deceleration distance.^21^
- *Six Minute Walk Test* - a measure of walking capacity and the primary outcome measure for this study. Participants are instructed to walk as far as they can in 6 minutes.^25^ Participants walk along a marked pathway required to be at least 20 meters long. Prior to starting the test, the participant is informed they may stop and rest in a standing or seated position as needed at any point, but that the timer does not stop.
- *Functional Ambulation Category* – a measure of walking independence, which is rated based on walking performance during the 10-meter and 6-minute walk tests.^26^

##### Self-reported measures

The following measures are completed by participants during the PRE, 4WK, 8WK, POST, and 3moPOST testing visit, with a blinded tester or non-treating study team member as needed:

- *Aerobic exercise outside of the study* – participants report the average number of times per week over the last month in which they performed aerobic exercise outside the study, the type of exercise performed (seated/walking), and whether exercise is usually vigorous enough to cause sweating (yes/no).
- *EuroQol 5-Dimension 5-Level* - a 5-item questionnaire about quality of life as it relates to mobility, self-care, usual activities, pain/discomfort, anxiety/depression, and overall health.^27^
- *Activities-Specific Balance Confidence Scale* – a 16-item questionnaire that asks participants to rate their balance confidence during every-day tasks on a scale from 0 to 100%.^28^
- *PROMIS-Fatigue Scale version 8a* - an 8-item questionnaire that inquires about the participant’s fatigue over the past seven days.^29^ Responses are rated on a 5-point Likert scale from “not at all” to “very much.”
- *Behavioral Regulation in Exercise Questionnaire* – a 24-item questionnaire assessing why participants may or may not engage in exercise.^30^ Responses are rated on a 5-point Likert scale from “strongly disagree” to “strongly agree”.
- *Global Ratings of Change–* asks participants to rate perceived changes in ease of walking, walking endurance, energy level and fatigue, and daily amount of walking activity since beginning the study (not applicable during PRE-testing). Responses are rated on a 7-point Likert scale from “much worse” to “much better”.

The following measure is performed at the end of the first training session and the middle of each 4 week treatment block (sessions 1, 6, 18, and 30) by the treating therapist:

- *PACES-8 Questionnaire* – an 8-item questionnaire assessing enjoyment of the exercise just performed.^31^

##### Step activity monitoring

Enrolled participants undergo continuous step activity monitoring throughout study participation using a StepWatch 4 (Modus Health, Edmonds, WA, USA) calibrated on their nonparetic ankle. Monitoring is initiated at least 3 days prior to beginning the intervention and continues until the end of the 3-month follow-up (study completion). Training session dates and times are recorded in the Modus Health Clinical Care Clinic App to identify stepping activity occurring within training sessions. At the start of each training session, step activity is uploaded, and the participant is provided feedback about daily walking averages. After the first week of training, the participant is encouraged to try to increase walking activity if they are not experiencing soreness or fatigue issues. If they are having soreness or fatigue but it is not interfering with daily activities, the participant is encouraged to maintain current walking activity. In the event of substantial prolonged soreness or fatigue, suggestions are made to temporarily decrease walking activity. To allow for remote uploads of the step activity data during the 3-month follow-up period, participants are assisted as needed in downloading the Modus Health Companion App onto their smartphones near their last training session. If a participant does not have a smart device, one is provided. The Modus Companion App performs automatic daily uploads to the cloud for remote data collection and provides participants their daily stepcount should they choose to look in the app.

#### Graded exercise test

Prior to enrollment, all participants undergo a graded exercise test (GXT) with a 12-lead electrocardiogram (ECG) and respiratory gas collection using a metabolic cart (TrueOne 2400, ParvoMedics, Salt Lake City, UT, USA). For the GXT, participants walk on a motorized treadmill with an overhead fall protection harness system and handrail use. Resting vitals are recorded and ECG monitoring is continuous throughout the GXT with blood pressure recorded every 2 minutes. The GXT begins with 3 minutes of walking at a pre-calculated individualized starting speed using the formula: GXT start speed (mph) = [CGS at PRE, rounded to the nearest 0.1 m/s] * 2.692 - 0.983, with a minimum of 0.3 mph.

This formula was developed using GXT data from HIT-Stroke Trial 1 and the PROWALKS trial,^32^ with the goal of having an average PRE GXT duration around 7 minutes (see Supplemental Content ‘HST2-GXT-protocol-section.html’ for details). This target average duration is slightly shorter than commonly utilized ranges among adults without stroke (8-12 minutes), to accommodate post-stroke neuromuscular fatigue. For ease of implementation, CGS at PRE is rounded to the nearest 0.1 m/s for this calculation, to reduce the number of protocols that needed to be programmed into the computers running the GXT treadmills. The starting speed used at the PRE GXT is also used for that participant at the POST GXT.

After the first 3 minutes of the GXT, speed is increased by 0.1 mph every 30 seconds until the participant demonstrates one or more termination criteria. The incline on the treadmill remains at 0% unless a participant achieves a speed of 3.5 mph, at which time speed is held at 3.5 mph and incline is increased 0.5% every 30 seconds until the participant demonstrates one or more termination criteria.^33^ Test termination criteria include the following: participant’s request to stop, drifting back on treadmill and being unable to recover, gait instability judged to pose an imminent safety risk by the testing therapist, and other stop criteria according to the American College of Sports Medicine guidelines.^9^ A physician or medical monitor reviews the ECG recording from the GXT to assess eligibility.

### Allocation

Participants are randomized 1:1 to either HIIT or MAT, with permuted block randomization stratified by site and by baseline walking speed (<0.4, ≥0.4 m/s), to help ensure groups are balanced within sites and on this prognostic factor.^15^ Eligible participants are randomized at the beginning of the first treatment session using the Research Electronic Data Capture (REDCap)^34^ randomization module. The study statistician who computer-generated the randomization sequence using SAS 9.4 and uploaded it to REDCap is the only person who has access to view it and has no interaction with study participants (e.g., not involved with recruitment or enrollment). Randomization cannot be triggered until eligibility is confirmed and is irreversible.

### Intervention features shared by both training protocols

#### Training volume and modes

The attendance goal for each group is 36 training sessions with a target frequency of 3x/week for 12 weeks, allowing up to one extra week for makeup sessions within each 12-session treatment block. For both groups, each training visit involves 45 minutes of walking exercise that consists of a 3-minute warm up of overground walking, 10 minutes of overground walking, 20 minutes of treadmill walking, 10 minutes of overground walking, and a 2-minute cool down of overground walking. During training, participants use their customary orthotic devices for all walking. Assistive devices for overground walking are selected based on which device enables achievement of the intervention intensity goal (fastest speed for HIIT; target HR for MAT). All training visits are performed by a PT to provide appropriate guarding for safety, and an overhead safety harness system is used for all treadmill training for fall prevention. No physical assistance or cueing is provided to change the participant’s gait pattern.

#### Heart rate and blood pressure monitoring

Participant heart rate is continuously monitored during training using either an optical heart rate sensor on the proximal forearm (e.g. Polar OH1+) or, if needed, an ECG sensor on the chest (e.g. Polar H10). The heart rate sensor is Bluetooth connected to an application (Fitdigits iCardio) on an Apple iPhone that can be worn on the forearm of the treating therapist. Target heart rate calculations are based on age-predicted maximal HR (206.9 – [0.67 x age]).^35^ For participants on beta blockers, maximal HR is predicted using: 164 – (0.7 x age).^36^ Blood pressure is measured at the start and end of each visit.

#### Session stop criteria

A session would be terminated if any of the following occur: (1) Signs of poor perfusion (e.g. cyanosis, pallor), (2) Drop in systolic blood pressure ≥10mmHg below the resting level from that day despite an increase in workload, (3) Hypertensive response with systolic blood pressure >240 or diastolic blood pressure >110, (4) New onset of significant nervous system symptoms (e.g. ataxia, dizziness, near syncope) or claudication pain, (5) Chest pain or angina, (6) Severe fatigue or shortness of breath in excess of what would be expected from exercise (e.g. shortness of breath not abating with rest), or (7) A serious injury (e.g. ankle sprain, fall with suspected fracture).

#### Additional challenges to increase intensity

Additional challenges can be added after the 6^th^ treatment visit if participants are not reaching intervention goals (e.g. speed plateau for HIIT, not reaching HR training target for either protocol) or if treadmill training speed has reached 3.5 mph. The additional challenge applied during overground training is a weighted sled drag with the sled anchor attached at waist level using an elastic band. Additional challenges during treadmill training include adding treadmill incline or resisted forward progression using an anchored bungee resistance band applied to the participant at waist level. The type of challenge and the amount of resistance are titrated by the treating therapist with the objective of best achieving training goals.

#### Training data collection

Overground training speed is measured with a stopwatch or automated speed gaits (VALD SmartSpeed Dash) as many times as possible during a bout and the peak speed without any additional challenge is recorded. Peak treadmill training speed without any additional challenge is recorded from the treadmill display during each treadmill bout. Participants report their rating of perceived exertion for the session using the 6-20 Borg scale^37^ at the end of every session, and step activity within the session is marked within the StepWatch Clinical Care App. Every 3 sessions, blood lactate measures, using a fingerstick, are taken following the treadmill training bout.

#### High-intensity interval training (HIIT) protocol

Individuals randomized to the HIIT group perform repeated 30 second bursts of walking at maximum safe speed, alternated with 30 to 60 second rest breaks. During overground HIIT, the first 2 rest breaks are 60 seconds and the remaining 7 rest breaks are 30 seconds, resulting in up to 9 total bursts during each overground bout. Speed is maximized by using visual targets to provide feedback on the distance covered during each burst, with encouragement to increase distance each burst. During treadmill HIIT, the first 3 rest breaks are 60 seconds and the remaining 15 rest breaks are 30 seconds, resulting in up to 18 total bursts. Fastest possible speed is the primary intensity target for HIIT. Speed is progressed to the participant’s fastest possible speed that does not require physical assistance and does not result in backwards drifting on the treadmill, gait instability posing imminent safety risk, toe drag into mid-swing, or excessive joint instability (e.g. ankle inversion, knee hyperextension) with risk of harm (e.g. near ankle sprain, knee snapping back forcefully). Fastest possible speed is the primary intensity target. A secondary goal is achieving an average HR of ∼70% HRR each session, with an expected range from 60-85% HRR.

#### Moderate-intensity aerobic training (MAT) protocol

Individuals randomized to the MAT group perform continuous walking during both overground and treadmill training. Training heart rate is the primary intensity target for MAT. Speed is continuously adjusted to maintain the following target HR ranges ± 5% HRR: training sessions 1-6: 40% HRR; training sessions 7-12: 45% HRR; training sessions 13-18: 50% HRR; training sessions 19-36: 55% HRR. During overground MAT, participants are instructed to increase or decrease their walking speed to maintain the desired HR. During treadmill MAT, speed is adjusted by the treating therapist to maintain the desired HR. Emphasis is placed on maintaining HR below 60% HRR since this is the threshold for vigorous intensity.^9^

### Safety monitoring

Safety monitoring includes active surveillance for AEs, with participant queries at the beginning and end of each study visit and direct observation for AEs throughout each study visit. The study team member completing the AE form provides a description of the event, its severity, its timing relative to study testing and/or intervention procedures, any possible alternative causes or contributing factors, any AE-related interventions (e.g., pain medicine), any follow-up, and if/when the event is resolved. A blinded centralized physician then adjudicates AEs for final categorization. Serious AE(s) include death, life threatening events, events that require inpatient hospitalization, or events that cause a persistent or significant disability/incapacity. AE(s) of particular interest are pain, fatigue, nausea, lightheadedness, cardiac disorder, fall, and injury.

### Data management

This study uses an electronic REDCap database hosted at Cincinnati Children’s Hospital Medical Center for direct electronic data entry and data management. The REDCap database includes automated calculations, prompts to increase protocol fidelity, and checks for impossible values or missing data. Additional data collection includes StepWatch activity data, iCardio continuous HR data, and SmartSpeed Dash overground training speed data, which are uploaded and stored on secure online servers.

### Standardization of procedures

All screening, assessment and intervention procedures are standardized, and study personnel complete a competency-based training program to assure that implementation is consistent across sites. All personnel are provided with the manual of operating procedures to review and complete video-based training modules related to their study roles. Personnel training also involves hands-on practice of study procedures with mock participants, which includes the use of all necessary equipment, participant monitoring, and real-time direct electronic data entry. Screening, testing and treating therapists are then certified through competency assessments for their study role(s). The multi-site team is also in frequent communication and meets regularly to ensure a consistent and standardized approach to any issues that arise during the trial.

### Statistical analysis

#### Specific aim 1

Determine the optimal locomotor training intensity for safely eliciting meaningful and sustained improvements in walking capacity among chronic stroke survivors.

##### Hypothesis 1a

Compared with MAT, HIIT will elicit meaningfully greater improvement in walking capacity, as measured by a 6MWD mean change difference significantly greater than +14 meters^38^ from PRE to POST.

##### Hypothesis 1b

Differences in 6MWD improvement between the treatment groups will be sustained 3 months after treatment completion (3moPOST). In other words, HIIT will elicit significantly greater 6MWD improvement than MAT from PRE to 3moPOST.

##### Hypothesis 1c

There will be no significant differences in the relative odds of serious adverse events between HIIT and MAT.

##### Analysis for hypotheses 1a and 1b

Testing for hypotheses 1a and 1b will use a general linear model with 6MWD as the dependent variable and fixed effects for [treatment group (HIIT, MAT)], [testing time point (PRE, 4WK, 8WK, POST, 3moPOST)], [group x time], [baseline walking limitation severity (<0.4 m/s, ≥0.4 m/s)], [baseline walking limitation severity x time], [study site] and [study site x time], with unconstrained variance & covariance between repeated testing time points within the same participant. Time will be modeled as a categorical factor (i.e. analysis of response profiles), to avoid making assumptions about the time course of changes. If there is a substantial sample size imbalance between sites, the [study site] term will be dropped from the model, to avoid giving excessive weight to a small number of participants at the lowest enrollment site(s). A ‘substantial imbalance’ will be considered present if the site with the highest enrollment has >20% more enrolled participants than the site with the smallest enrollment.

Hypothesis 1a will be tested by the [group x time] effect from PRE to POST, against a null hypothesis of a +14-meter mean change difference for HIIT vs. MAT, using a one-sided minimal effects test.^3^ If the null hypothesis is not rejected, a two-sided test against a null hypothesis of zero mean change difference will be performed to assess the possibility of statistically significant but not clinically meaningful between group differences, or significant and meaningful differences favoring MAT. Hypothesis 1b will be tested by the [group x time] effect from PRE to 3moPOST, against a null hypothesis of zero mean change difference, using a two-sided test. Both hypotheses will be tested at the .05 significance level since they are addressing distinct scientific questions. Missing data will be handled with the method of maximum likelihood, which assumes data are missing at random (MAR). A sensitivity analysis will assess whether conclusions depend on the MAR assumption. Secondary outcomes will also be tested using the same modeling, with false discovery rate (FDR) correction across both time points and all measures, with the Benjamini-Hochberg procedure.^39^

To preliminarily assess for differences in optimal intensity between persons with mild/moderate vs severe baseline walking limitations,^15,40^ we will also add a [treatment group x baseline walking limitation severity x time] interaction term to the primary model, acknowledging that this analysis will not be well powered. There is no strong evidence or compelling reason to suspect that the optimal locomotor training intensity depends on other baseline covariates (e.g. sex). However, we will still preliminarily assess this possibility by testing for [treatment group x covariate x time] interactions, using a separate model for each covariate (while also including the lower order terms involving the covariate in the model). If the interaction is not significant, differences in overall training responsiveness will be tested by the [covariate x time] interactions, after dropping the [treatment group x covariate x time] interaction term from the model. The significance level will be FDR corrected across cofactors, except when testing the prespecified stratification factor (baseline walking limitation severity) and the following baseline covariates, which are being prespecified based on a prior analysis: Fugl-Meyer lower limb motor function score, Brief Balance Evaluation Systems Test score, Activities-specific Balance Confidence Scale score, age, participant report of pain-limited walking duration, and participant report of prior walking exercise >2 days/week.^41^

##### Analysis for hypothesis 1c

To test hypothesis 1c, post-randomization adverse events (AE) will be compared between treatment groups with logistic regression. Separate models will be tested for overall AE(s) and for each AE categorization, using the number of participants with AE(s) in that category as the dependent variable and fixed effects for [treatment group], [study site] and [baseline walking limitation severity (<0.4, 0.4-1.0 m/s)]. As described above, the [study site] term will be dropped from the model if there is a substantial sample size imbalance between the sites. If only one group has participant(s) with AE(s) in a particular category, continuity correction will add 0.5 participants with and without an AE to each group to permit odds ratio calculation. In this case, it is not possible to adjust for study site or baseline walking limitation severity.

#### Specific aim 2

Determine whether vigorous training intensity is particularly critical for patients with severe walking speed limitations.

##### Hypothesis 2a

Compared with MAT, HIIT will elicit significantly greater improvements in walking capacity for persons with severe baseline walking limitations (speed <0.4 m/s) and for persons with mild/moderate baseline walking limitations (speed ≥0.4 m/s), as measured by mean 6MWD changes from PRE to POST.

##### Hypothesis 2b

Among participants with **severe** baseline walking limitations (speed <0.4 m/s), MAT will not elicit meaningful improvement in walking capacity from PRE to POST, as indicated by a mean 6MWD change (and upper 95% confidence limit) <20 meters.^42^

##### Hypothesis 2c

Among participants with **severe** baseline walking limitations (speed <0.4 m/s), HIIT will elicit meaningful improvement in walking capacity from PRE to POST, as indicated by a mean 6MWD change (and lower 95% confidence limit) ≥20 meters.^42^

##### Hypothesis 2d

Among participants with **mild/moderate** baseline walking limitations (speed 0.4-1.0 m/s), both HIIT and MAT will elicit meaningful improvement in walking capacity from PRE to POST, as indicated by mean 6MWD changes (and lower 95% confidence limits) ≥20 meters.^42^

##### Analysis for Aim 2

A somewhat larger and more typical meaningful change threshold is being applied for the within-group 6MWD changes in hypotheses 2b-2d (+20 meters) versus the smallest between-group difference of interest for hypothesis 1a (+14 meters). The within-group change threshold of +20 meters is still on the lower end of typically used 6MWD change thresholds, which usually range from +20 to +50 meters.^38,42^

Testing for hypotheses 2a-2d will use the same general linear model as hypotheses 1a-1b above, but with an added [baseline walking limitation severity x treatment group x time] interaction. Variances will also not be constrained to be the same between baseline walking limitation severity subgroups or treatment groups. Hypothesis 2a will be tested by two-sided time contrasts from PRE to POST between treatment groups, within each baseline walking limitation severity subgroup, at the .05 significance level. Hypotheses 2b-2d will be tested by one-sided time contrasts from PRE to POST against a null hypothesis of +20 meters within the relevant baseline walking limitation severity subgroups and treatment groups. These nested contrasts will also use the .05 significance level but will only be tested if the corresponding between-group difference in mean change from hypothesis 2a is statistically significant (i.e. hierarchical testing). Secondary outcomes will be tested with FDR control across all measures.^39^ As in Aim 1, the primary analysis will handle any missing data with the method of maximum likelihood (under the MAR assumption), and the same MAR sensitivity analysis from Aim 1 will be performed. We will also test for baseline covariate effects using the same methods described above for Aim 1. See ClinicalTrials.gov record NCT06268040 for the full analysis plan.

### Sample size determination

Sample size was calculated to provide sufficient power for testing hypothesis 1a and was also determined to provide sufficient power for testing the remaining hypotheses. A simulation was conducted using 6MWD population parameters that were estimated using data from HIT-Stroke Trial 1 (N=55).^14^ MAT means and residual (co)variances were taken directly from primary analysis results.^14^ HIIT means were conservatively set at 80% of the observed mean change differences from MAT (i.e. 80% of +15, +29 and +44 meters at 4WK, 8WK and POST, respectively). Percentages of missing data were set at the observed values after excluding missingness due to COVID-19 shutdown and conservatively rounding up to the nearest whole percent. For each potential sample size, 1,000 random samples were drawn from a multivariate normal distribution with these population parameters. The proposed minimal effects test for hypothesis 1a was performed in each sample, and the proportion of samples where the null hypothesis was rejected was taken as the power for each sample size. The power curve was then smoothed using LOESS and the smallest sample size with ≥80% power was identified. This simulation estimated a total needed enrollment of 156 participants (78 per group). See ClinicalTrials.gov record NCT06268040 for full details of the power analysis.

### Data monitoring

Intervention fidelity is monitored using intervention data including treadmill training speed in relation to overground training speed as a measure of neuromotor intensity, blood lactate values as a measure of anaerobic intensity, mean and max HR as a measure of aerobic intensity, and the number of sessions attended. To minimize risk of bias, outcome data are only monitored for missing or implausible values using automated processes within REDCap, without regard to randomization group.

AEs and protocol deviations are reported by study staff to the site PIs on a regular basis and are discussed during multi-site conference calls. All identified AEs and protocol deviations are also reported to the UC Institutional Review Board (IRB) and an independent Data and Safety Monitoring Board (DSMB) annually, regardless of suspected causal relationship to the study treatment.

The DSMB acts in an advisory capacity to the IRB to monitor participant safety, data quality and evaluate the progress of the study. It consists of at least three members with combined expertise in stroke rehabilitation, clinical trials and biostatistics, who do not have paid appointments at the study institutions, and do not have a collaborative or mentoring relationship with the study investigators. The DSMB will meet each year of the study, either in person or via teleconference, and additional meetings can be scheduled if needed. The DSMB evaluates participant accrual & retention, baseline characteristics, protocol compliance and AE data. Clinical outcome data can also be provided if requested by the DSMB to weigh the risks against the benefits of study participation. However, no interim analyses are planned. The DSMB assesses the risks of study participation for all participants and provides a written report of their analysis and recommendation as to whether the study should continue, whether modifications to the study are needed or if the study should be terminated. DSMB meetings include open sessions where the DSMB may discuss any issues with the study team and closed sessions where the DSMB alone decides on its recommendations.

## DISCUSSION

This study will provide more precise estimation of the relative benefits and risks of vigorous vs. moderate intensity locomotor training in chronic stroke, to determine the optimal intensity for safely eliciting meaningful and sustained improvements in walking capacity. It will also more precisely estimate responsiveness to vigorous vs. moderate intensity locomotor training among individuals with severe walking limitations (CGS <0.4 m/s), to assess whether vigorous training intensity is particularly critical for this lower prognosis subgroup. As such, the results of this study are expected to provide more specific guidance for optimizing locomotor training intensity and walking outcomes in chronic stroke.

## Data Availability

The datasets generated and/or analyzed during this current study will be uploaded to NICHD data and specimen hub at the time of results publication.

## List of abbreviations

HRR: heart rate reserve
HIIT: high-intensity interval training
MAT: moderate-intensity aerobic training
6MWD: 6-minute walk distance
PRE: pre-testing
PT: physical therapist
4-WK: after 4 weeks of training
8-WK: after 8 weeks of training
POST: POST - the end of the 12-week training protocol
3moPOST: 3 months after the completion of the last training session
CGS: comfortable gait speed
UC: University of Cincinnati
UD: University of Delaware
KUMC: University of Kansas Medical Center
mph: miles per hour
ECG: electrocardiogram
GXT: graded exercise testing
HR: heart rate
REDCap: Research Electronic Data Capture
MAR: missing at random
FDR: false discovery rate
AE: adverse event
IRB: Institutional Review Board
DSMB: Data and Safety Monitoring Board

## Declarations

- Ethics approval and consent to participate, include the name of the ethics committee that approved the study: The University of Cincinnati Institutional Review Board Research approved this study on January 12, 2024. Trial Status: University of Cincinnati Institutional Review Board Research approved Protocol 2017-5325 version 7 was the initial approval for this study on January 12, 2024 and updated to version 8 on August 13, 2024. Recruitment Start Date: February 9, 2024; Approximate recruitment completion date: April, 2028
- Consent for publication – Not applicable
- Availability of data and materials – The datasets generated and/or analyzed during this current study will be uploaded to NICHD data and specimen hub at the time of results publication.
- Competing interests - The authors declare that they have no competing interests.
- Funding: Eunice Kennedy Shriver National Institute of Child Health and Human Development (NICHD) R01HD093694

## Acknowledgements

None

## Notes

### Competing Interest Statement

The authors have declared no competing interest.

### Clinical Trial

NCT06268041

### Author Declarations

The University of Cincinnati Institutional Review Board Research approved this study on January 12, 2024.

